# An Agent-Based Model to assess COVID-19 spread and health systems burden in Telangana state, India

**DOI:** 10.1101/2020.10.03.20206177

**Authors:** Narassima M S, Guru Rajesh Jammy, Sankarshana A, Rashmi Pant, Anbuudayasankar S P, Lincoln Choudhury, Vijay Yeldandi, Shubham Singh, Denny John

## Abstract

**Objectives:** To assess the transmission dynamics and the health systems’ burden of COVID-19 using an Agent Based Modeling (ABM) approach using a synthetic population.

**Study design:** The study used a synthetic population with 31,738,240 agents representing 90.67 percent of the overall population of Telangana state, India as per 2011 Census of India. Lockdown phases as per Indian scenario considering the effects of post-lockdown, use of control measures and immunity on secondary infections were studied.

**Methods:** The counts of people in different health states were measured separately for each district of Telangana. The model was run for 365 days and six scenarios with varying proportions of people using control measures (100%, 75% and 50%) and varying immunity periods (90 and 180 days). Sensitivity Analysis has been done for two districts to compare the change in transmission dynamics when incubation period and asymptomatic proportion are changed.

**Results:** Results indicate that the peak values were attained soon after the lockdown was lifted. The risk estimates indicate that protection factor values are higher when more proportion of people adopt control measures.

**Conclusions:** ABM approach helps to analyze grassroot details compared to compartmental models. Risk estimates allow the policymakers to determine the protection offered, its strength and percentage of population shielded by use of control measures.

## 1. Introduction

On Jan 30, 2020, India reported its first infection of Covid-19 (SARS-CoV-2) during when WHO categorized Covid-19 to be “Public Health Emergency of International Concern” (PHEIC), owing to the serious impacts that could be caused by the infection [16; 53]. The world has witnessed an enormous outbreak of the epidemic with 34,503,272 infections and 1,027,138 deaths across 213 nations worldwide, as on September 30, 2020 resulting in a global health crisis [77]. In India, total infections reported are 6,310,267 with 940,643 active cases, 5,270,007 recoveries and 98,708 deaths, till September 30 2020 [17]. Countries like India, with higher population densities have a greater concern owing to the influx of infections [38]. The severity of infection and recovery varies from case to case, majorly governed by some parameters such as comorbidities, age, exposure to virus particles, air pollution, etc. [19; 44]. In addition, majority of the infections being asymptomatic raise a serious threat as they remain untraceable and continue to transmit the infection [19; 44; 72]. There is a lot of work being done by researchers, policymakers, healthcare professionals across various disciplines to rapidly eradicate the spread of the infection [26; 52].

Several studies on infectious diseases dealing with containment of diseases such as tuberculosis [55], measles [23], etc., operationalizing antiviral prophylaxis to control H5N1 influenza and distancing [25], strategizing evacuations in the case of airborne infections [24], establishing vaccination techniques for smallpox [34], influenza [15], etc. Presently, ABMs have been developed to study COVID-19 related scenarios like the effectiveness of imposing lockdowns [40; 51; 68; 71], post-lockdown control strategies [37], insulation of vulnerable population [37; 36], direct and indirect transmission (via viral particles in air) [40], effect of control measures like distancing and face mask [37; 43], transmission based on viral-load [43], population intelligence [68], contact tracing initiatives [43; 68], contacts based on schedule and locations [21; 37; 40], etc.

Most studies from India on COVID-19 have largely followed compartmental approach adhering to either the basic Susceptible (S), Infective (I) and Recovered (R) model or its variations that include additional states. These include, the Susceptible (S), Exposed (E), Symptomatic (I), Purely Asymptomatic (P), Hospitalized or Quarantined (H), Recovered (R) and Deceased (D) (SIPHERD) [50], Susceptible (S), Exposed (E), Infective (I) and Recovered (R) (SEIR) [13; 66; 74; 78], analytical models [2; 67].

Simulation models map the real-world behaviour through a set of rules, with the defined level accuracy, subject to constraints [6; 7]. These models eliminate the investment of cost, time and associated risks [7; 48]. Complex problems involving dynamicity are much effectively handled by simulations [30]. Three widely used simulation approaches are System Dynamics (SD), Discrete Event Simulation (DES) and Agent Based Modeling (ABM). SD and DES provide only collective measures whereas ABM holds granular details of individual agents [11; 69]. ABM allows modelers to define parameters uniquely to agents [12; 30; 69].

ABMs are resultant of advancements in science and technology and the ability of systems to handle complexities [30; 49]. In recent times, these models are sought by researchers across various sectors [33; 31; 32]. ABM incorporates a bottom-up approach wherein the behavior of agents cumulate to the behaviour of the system [12; 27]. This has attracted the public health researchers and practitioners as they can observe the actions of individuals and clearly apprehend the population dynamics better [6; 60]. However, the computational capabilities majorly govern the potential of ABMs [49]. In this paper, we have used ABM approach to analyse outbreak and health systems burden on COVID-19 in Telangana state. Multiple NPIs have been considered simultaneously as enforced by the Swiss Cheese model so that each layer curtails the spread of infection to some extent [29; 56]. This has been in use by hospitals to establish multiple defense strategies [56] and also by policymakers to impose multiple interventions [29].

## 2. Methodology

### 2.1. Research design

The current study employs an Agent Based Modeling (ABM) approach to analyze the outbreak and health systems burden for Covid-19 with the synthetic population of Telangana state (Table 1). The model was coded in python using PyCharm (Version: 2020.1.3). Python being an Object-Oriented Programming (OOP) language was chosen to code the model as OOP are more suitable for ABM [4]. The model was run for 365 days considering the lockdown phases as per Indian scenario (Table 3). The study involves agent creation, establishing a contact network, creating a disease model and initializing the model. This is followed by running the model, extracting and analyzing the results and providing useful interpretations. Public health policies and recommendations are underpinned based on the estimations of the mathematical models devised [9; 63]. The code, scenarios, parameters and scope of the model are all made transparent in the present study and adheres to ethical good practices in modelling and International Society for Pharmacoeconomics and Outcomes Research (ISPOR-SMDM) Modeling Good Research Practices [9; 10; 63].

**Table 1:**
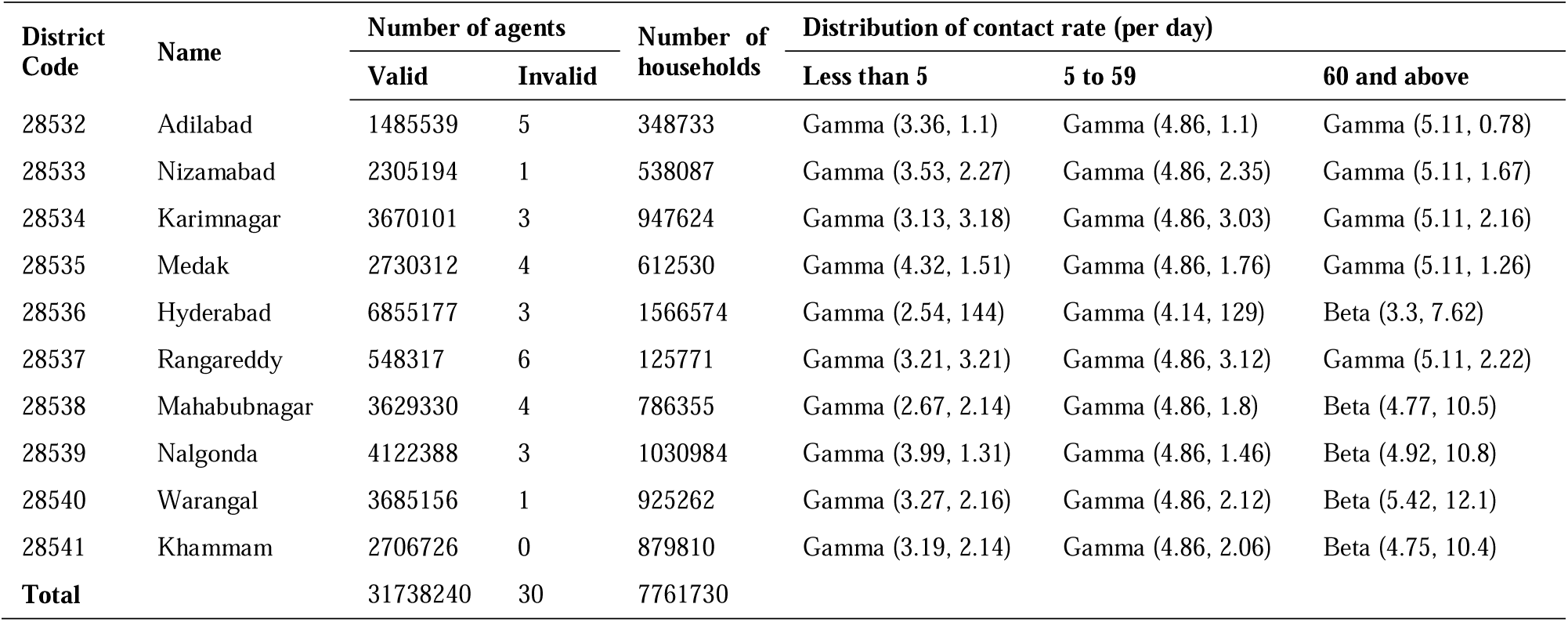
Number and contact rate distributions of agents

| District Code | Name | Number of agents |  | Number of households | Distribution of contact rate (per day) |  |  |
| --- | --- | --- | --- | --- | --- | --- | --- |
|  |  | Valid | Invalid |  | Less than 5 | 5 to 59 | 60 and above |
| 28532 | Adilabad | 1485539 | 5 | 348733 | Gamma (3.36, 1.1) | Gamma (4.86, 1.1) | Gamma (5.11, 0.78) |
| 28533 | Nizamabad | 2305194 | 1 | 538087 | Gamma (3.53, 2.27) | Gamma (4.86, 2.35) | Gamma (5.11, 1.67) |
| 28534 | Karimnagar | 3670101 | 3 | 947624 | Gamma (3.13, 3.18) | Gamma (4.86, 3.03) | Gamma (5.11, 2.16) |
| 28535 | Medak | 2730312 | 4 | 612530 | Gamma (4.32, 1.51) | Gamma (4.86, 1.76) | Gamma (5.11, 1.26) |
| 28536 | Hyderabad | 6855177 | 3 | 1566574 | Gamma (2.54, 144) | Gamma (4.14, 129) | Beta (3.3, 7.62) |
| 28537 | Rangareddy | 548317 | 6 | 125771 | Gamma (3.21, 3.21) | Gamma (4.86, 3.12) | Gamma (5.11, 2.22) |
| 28538 | Mahabubnagar | 3629330 | 4 | 786355 | Gamma (2.67, 2.14) | Gamma (4.86, 1.8) | Beta (4.77, 10.5) |
| 28539 | Nalgonda | 4122388 | 3 | 1030984 | Gamma (3.99, 1.31) | Gamma (4.86, 1.46) | Beta (4.92, 10.8) |
| 28540 | Warangal | 3685156 | 1 | 925262 | Gamma (3.27, 2.16) | Gamma (4.86, 2.12) | Beta (5.42, 12.1) |
| 28541 | Khammam | 2706726 | 0 | 879810 | Gamma (3.19, 2.14) | Gamma (4.86, 2.06) | Beta (4.75, 10.4) |
| **Total** |  | 31738240 | 30 | 7761730 |  |  |  |

**Table 2:** Model parameters

| Parameters | <5 | 5-59 | >59 | References |
| --- | --- | --- | --- | --- |
| Number of contacts per day | Table 1 |  |  | [8; 46] |
| Probability of getting infected through contact | i) household/ closer circle: (3 to 10) percent<br>ii) other contacts: (1 to 5) percent |  |  | [5] |
| Proportion of people remaining asymptomatic | 0.8 |  |  | [22; 75] |
| Average incubation period (in days) | 5 |  |  | [12; 79] |
| Average treatment duration (in days) | 14 |  |  | [79] |
| Proportion of hospitalized cases in ICU | 0.11 |  |  | [3; 28] |
| Treatment duration in ICU (in days) | Triangular(7,8,9) |  |  | [28; 79] |
| Proportion of people moving from ICU to critical illness (Ventilator assistance) | 0.88 |  |  | [28] |
| Treatment duration in ventilator state (in days) | Triangular(5,7,12) |  |  | [79] |
| Time between symptom arrival and admission (in days) | 3 |  |  | [76] |
| Proportion of people who die | As per Indian statistics |  |  | [17] |
| Risk difference for use of control measures (percentage) | i) Mask: 10.2<br>ii) Distancing: 14.3 |  |  | [14] |

**Table 3:**
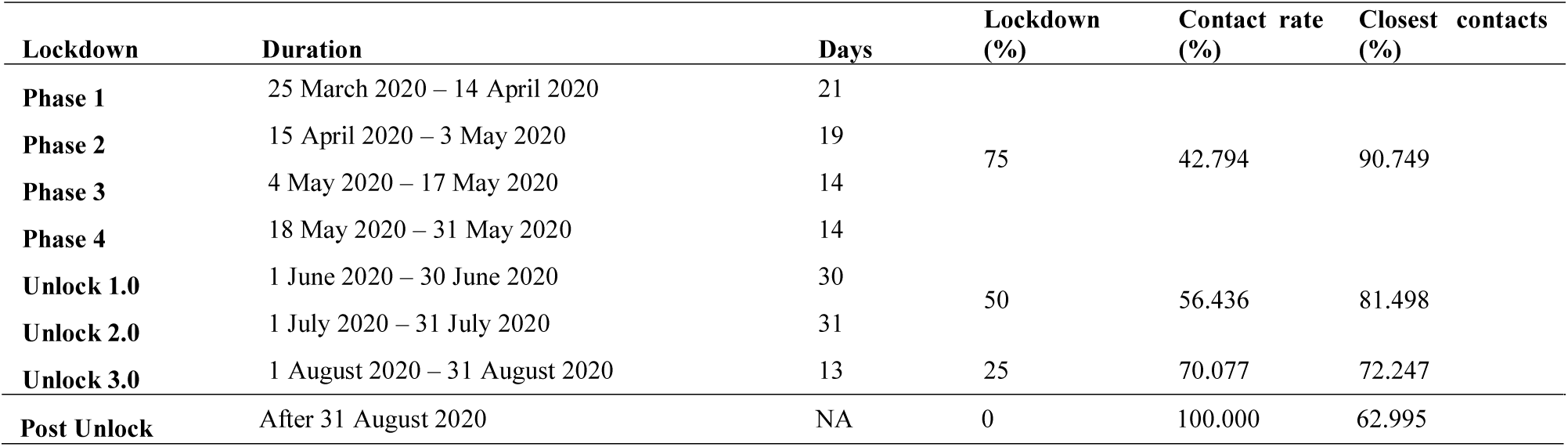
Lockdown phases

### 2.2. Agent creation

Synthetic population developed using the 2011 Census of India data for 31,738,270 people was used to generate agents. Agent attributes such as person ID, age, household ID, geocoordinates, and district code were assigned to each agent. During this process, 30 invalid records were skipped thereby making the valid number of agents to be 31,738,240 (Table 1). This count represents 90.67 percent (n=35,003,674) of the overall Telangana population as per 2011 Census [57; 70].

### 2.3. Contact network

Transmission of an infection is majorly governed by transmission rates and contact network. As per the WHO report on COVID-19 (16 to 24 Feb 2020), transmission rates were randomly varied from 3 to 10 percent for household and closer contacts and 1 to 5 percent for other contacts [5]. To establish varying contact rates for each district, a Density-Dependent contact rate was assumed [39; 58]. Kumar et al. (2018) determined the contact rates for close contact infections by considering the case of Ballabgarh, India [47]. The population density of Ballabgarh and those of the ten districts of Telangana chosen for the study were used to derive the contact rates for the ten districts. The ratio of the population density of Ballabgarh to that of each district would be the corresponding multiplication factor to scale up the number of contacts made by each person in Ballabgarh, as presented by Kumar et al. (2018) [39; 47; 58]. The distributions followed by the datasets consisting the contact rates of individuals of each district were determined using Arena’s ‘Input Analyzer’ tool (Arena 16.00.00002) (Table 1). The tool generates different distributions to which the input datasets could fit, with the associated errors [65].

### 2.4. Disease model

Disease model helps to represent the agent behaviour and trajectory of a disease using various health states [1]. Each agent can exist in any one of the states described by the state chart at a moment (figure 1). The transition between these states and the duration for which an agent remains in a state are defined as presented in table 2. During the simulation, agents interact based on the contact rates during which an infected agent transmits the infection to a healthy agent. Infected agents who are asymptomatic recover without treatment but continue to transmit the infection till they recover. Symptomatic individuals seek treatment after incubation period. They either recover or decease whilst in any of the treatment levels defined as admitted, ICU and ventilator.

**Figure 1:**
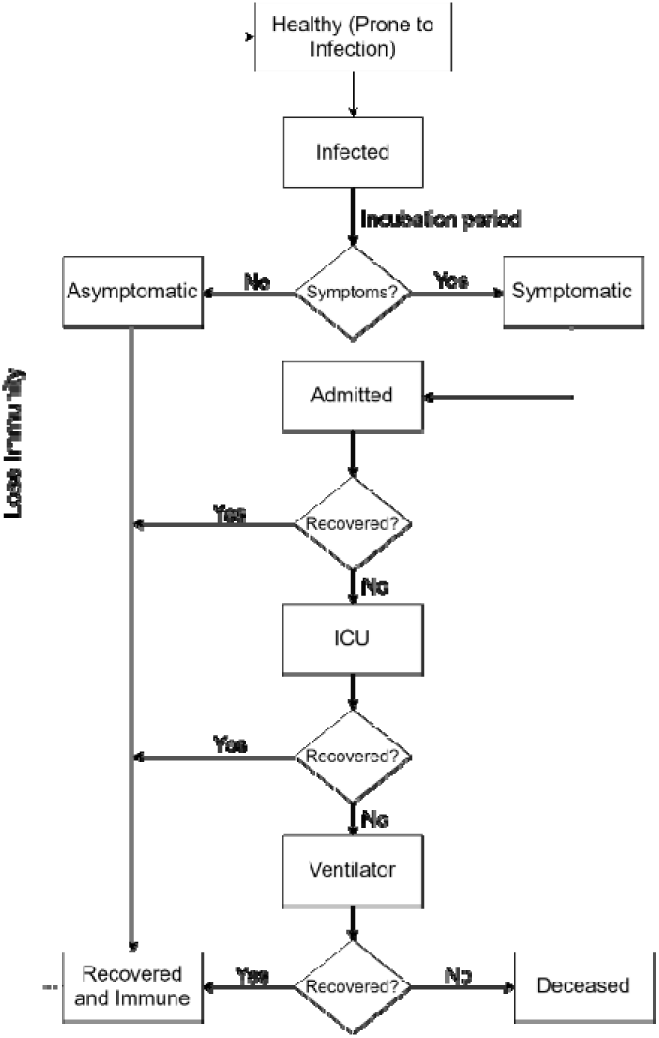
State Chart

### 2.5. Model initialization

Parameters that are required to drive the model were acquired from various sources including Models of Infectious Disease Agent Study (MIDAS) [54].

### 2.6. Model simulation

The model coded in Python was equipped with the input parameters and run for six different scenarios (table 4) [18]. The lockdown stringency was varied for various phases of lockdowns imposed. The proportion of contacts made by Indians based on locations were determined [61]. To bring in the effect of lockdown, contacts made by people at work and other places were reduced proportionately based on lockdown stringency, the contacts in home were maintained same and the school contacts were nullified.

**Table 4:**
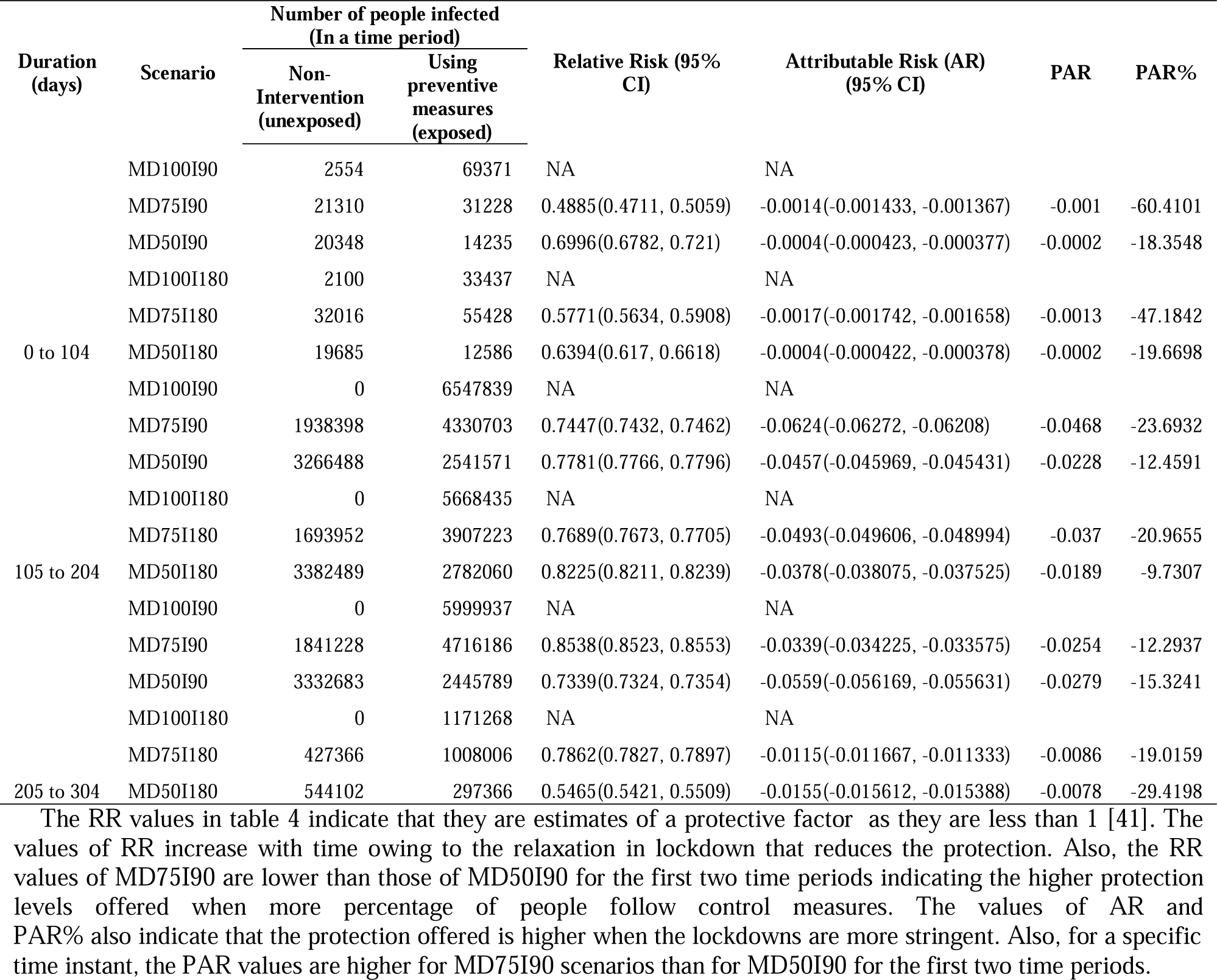
Risk estimations

## 3. Results of Simulation

The model was run for six different scenarios considering the various phases of lockdown imposed in India, control measures such as use of face mask and social distancing and the impact of immunity on the transmission of infection. The six scenarios would be referred to as MD100I90, MD75I90, MD50I90, MD100I180, MD75I180 and MD50I180 in subsequent sections. The number following ‘MD’ indicates the percentage of people following control measures and the number following ‘I’ indicates the days for which people remain immune after recovery. The graphs plotted represent the number of people in each health state for the entire state of Telangana. District-wise counts are provided in the supplementary excel.

It is clear from figures 2 a) and b) that the number of uninfected people decrease significantly once the lockdown has been completely lifted after 142 days. As the stringency of lockdown is reduced, people meet more people increasing the vulnerability of acquiring or transmitting infection. In both these figures, there is a visible increase in the number of uninfected people which indicates the transit of people from immune state to uninfected state i.e., loss of immunity/ prone to infection [62].

**Figure 2.**
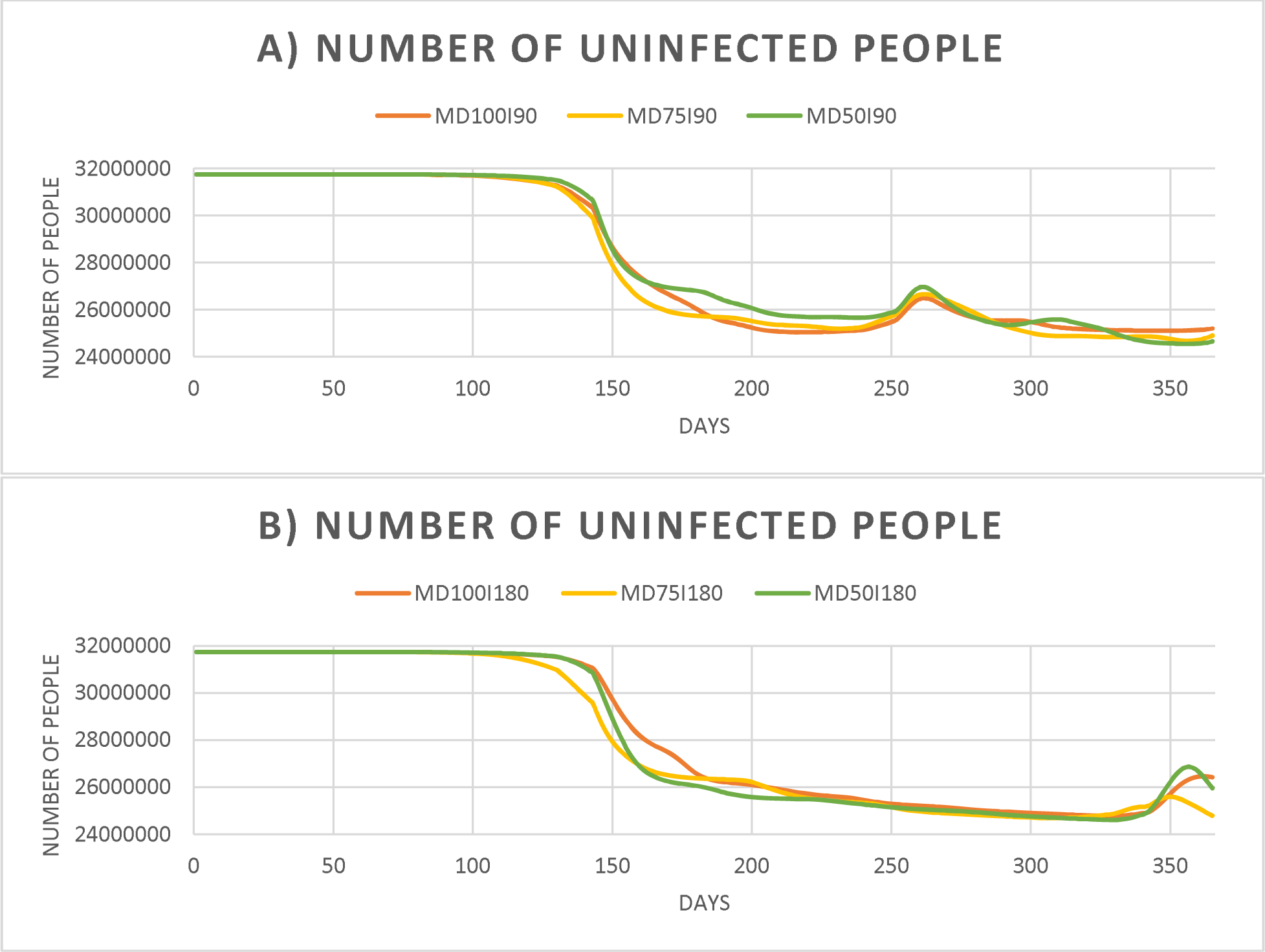
a): Number of Uninfected people (90 days immunity scenarios); b) Number of Uninfected people (180 days immunity scenarios)

In figures 3 a), b), c) and d), it is evident that the infection has begun to rise as the lockdown is lifted. The second spike in figures 3 a) and c) indicates the secondary infection which is because of the loss of immunity among recovered people. The same phenomena can be observed to have just begun at the end of one year in figures 3 b) and d). Though control measures do not curtail the spread of infection as effectively as lockdown, it offers a level of protection, reducing risk. This is evident from the above figures where the peak values are lesser for the scenarios in which higher proportion of people follow control measures.

**Figure 3.**
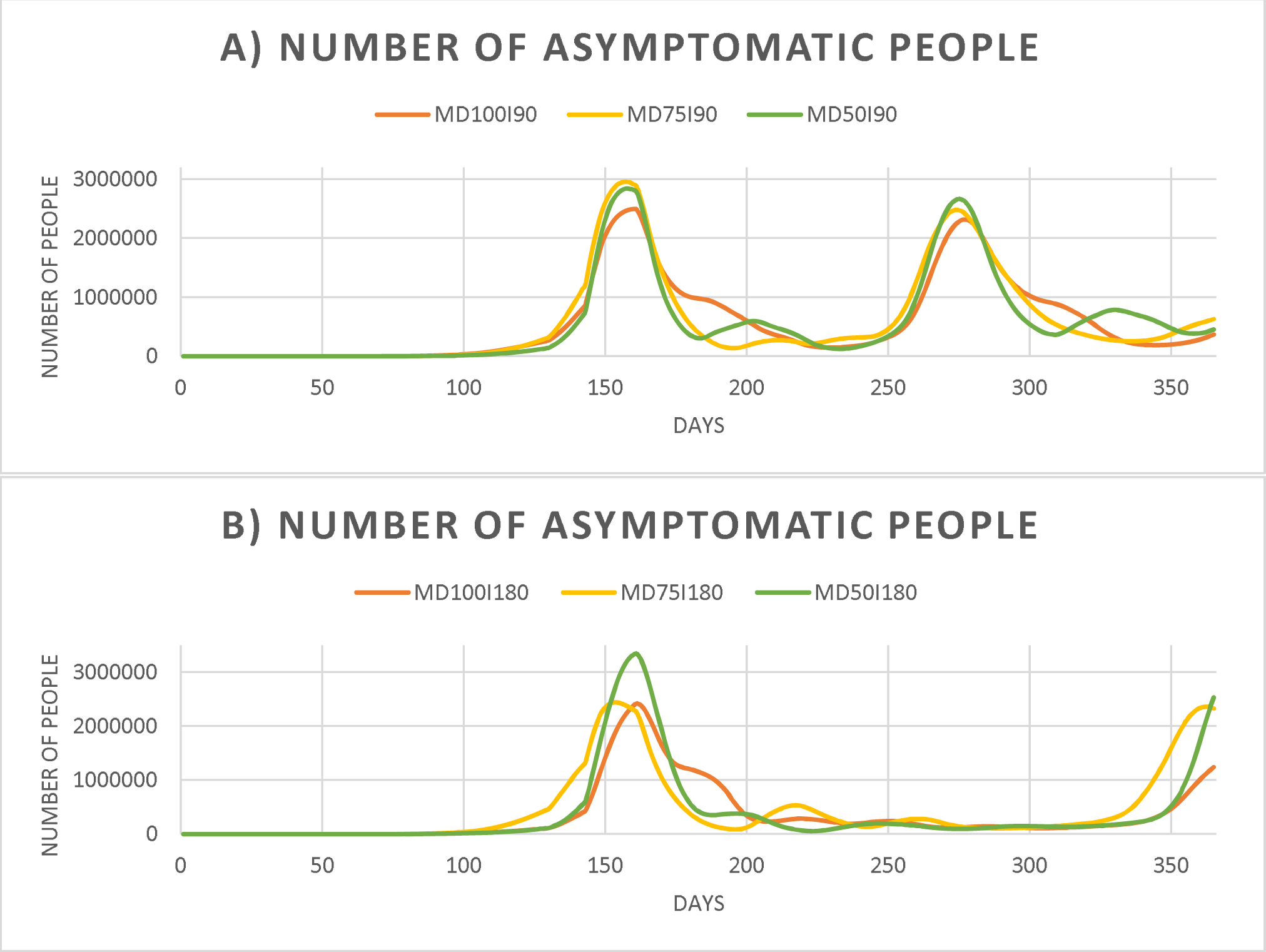
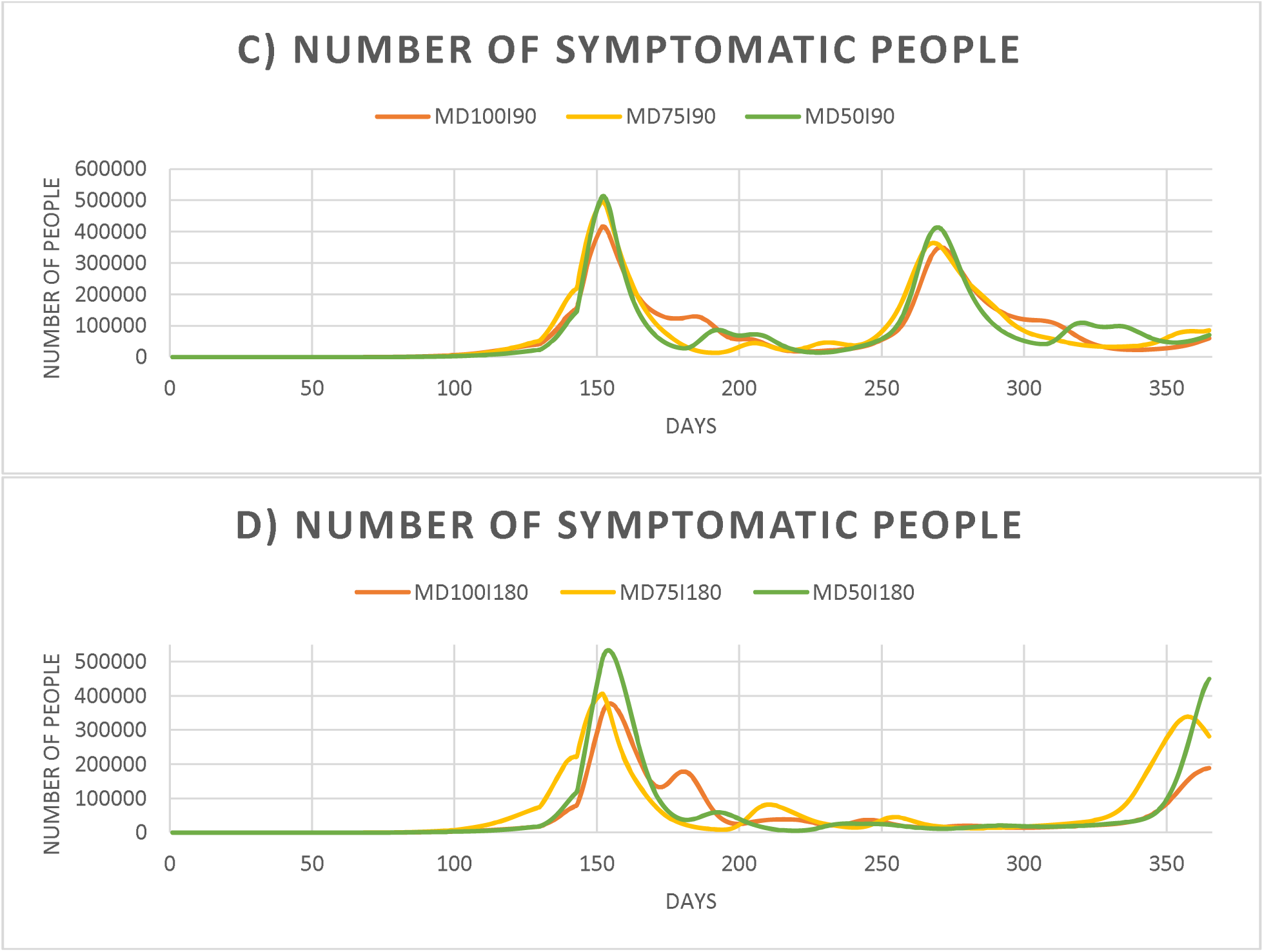
a): Number of Asymptomatic people (90 days immunity scenarios); b) Number of Asymptomatic people (180 days immunity scenarios); c): Number of Symptomatic people (90 days immunity scenarios); b) Number of Symptomatic people (180 days immunity scenarios)

From figures 4 a), b) c), d), e) and f), we can observe that there is always a second influx of admissions possible in absence of vaccination. These graphs provide information that could help policymakers, health care system and government to plan their capacity to accommodate the patients and make arrangements for intensive care and ventilators. Similar observations as in figure 3 are observed in terms of secondary infections post loss of immunity of people.

**Figure 4.**
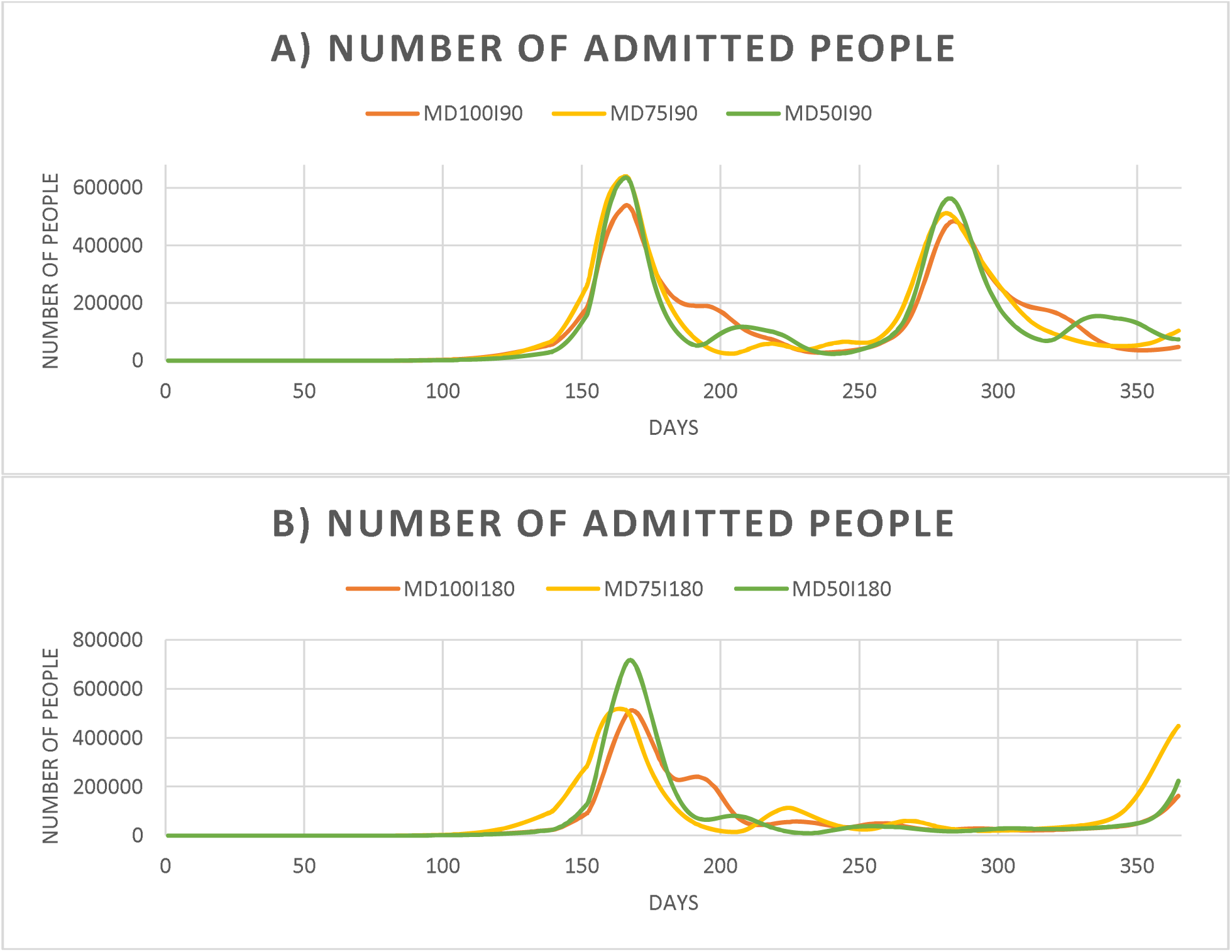
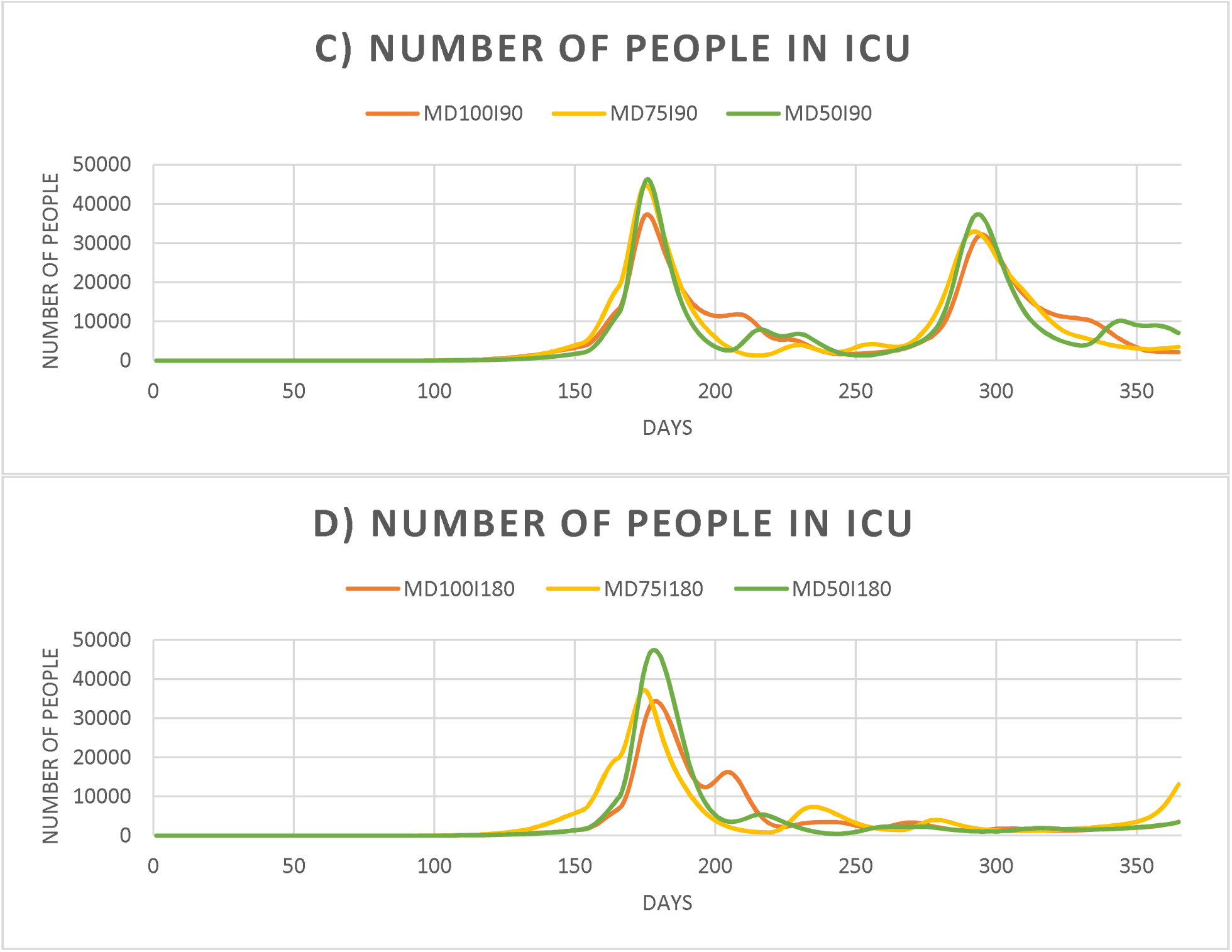
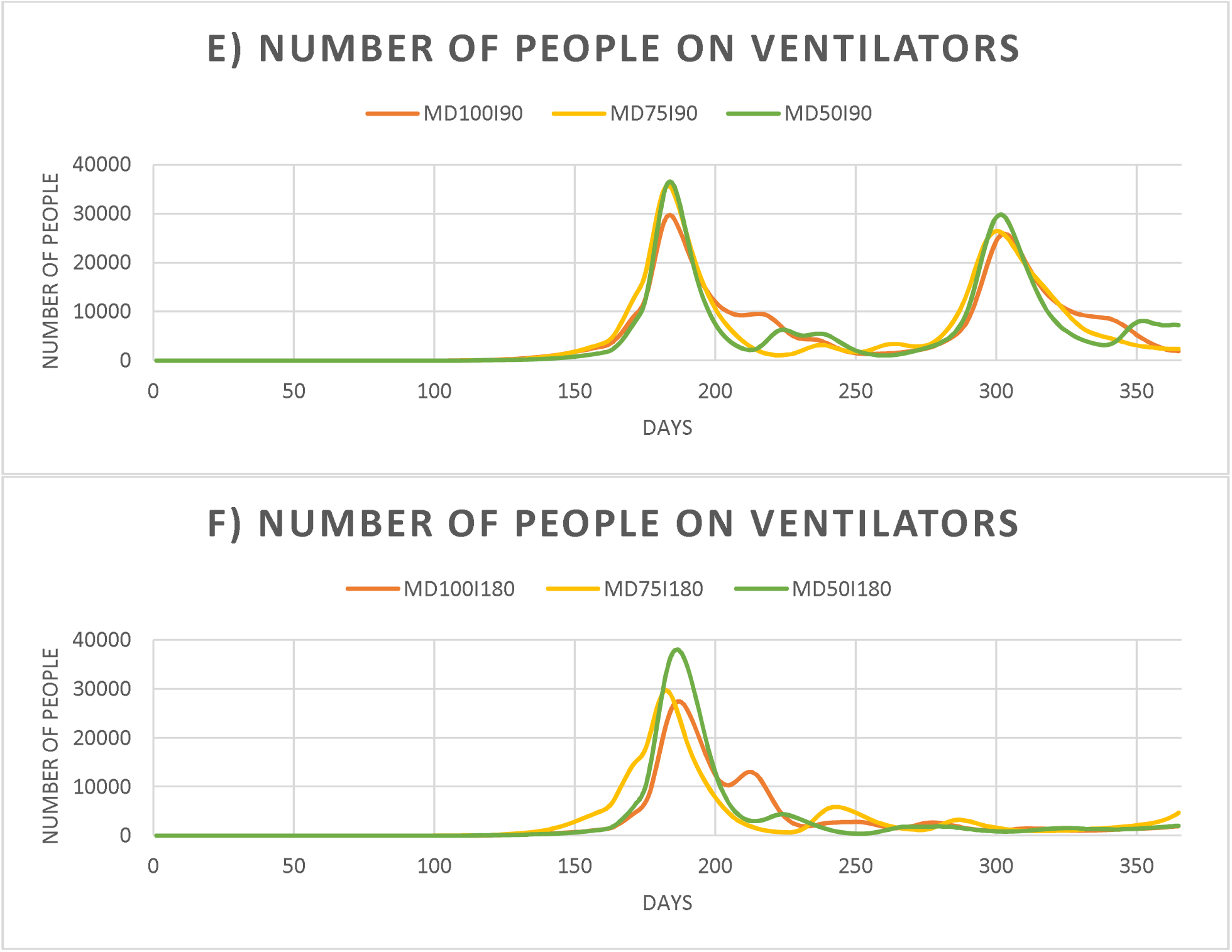
a): Number of Admitted people (90 days immunity scenarios); b) Number of Admitted people (180 days immunity scenarios); c): Number of people in ICU (90 days immunity scenarios); b) Number of people in ICU (180 days immunity scenarios); e) Number of people on Ventilators (90 days immunity scenarios); b) Number of people on Ventilators (180 days immunity scenarios)

Figures 5 a) and b) represent the number of deceased people. The spike in this number is observed post-lockdown and an additional spike is observed in 5 a) indicating the second influx of infections. This can be directly related to the loss of immunity that is seen in figures 5 c) and d). Longer immunity offers the health care systems more time to devise vaccination strategies and capacity planning.

**Figure 5.**
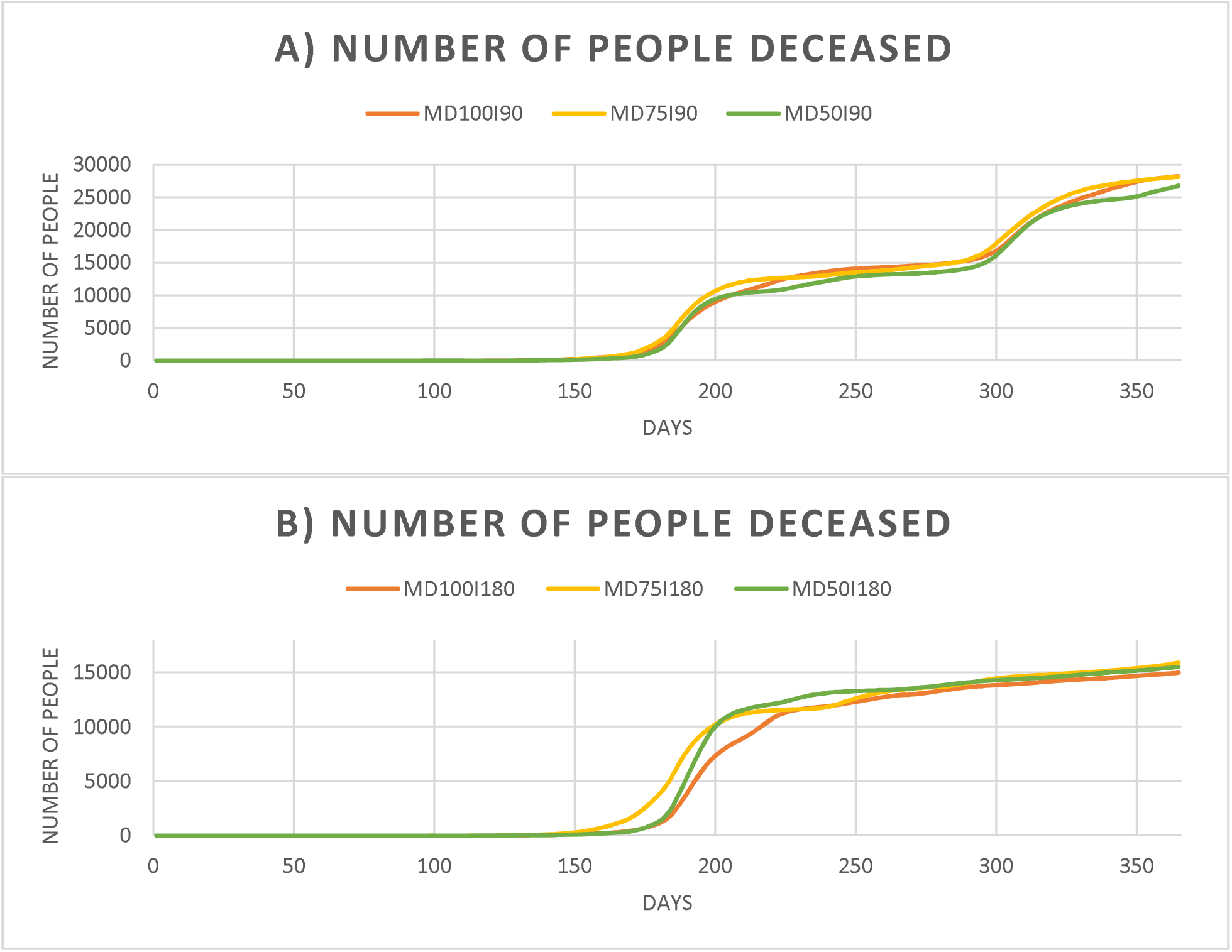
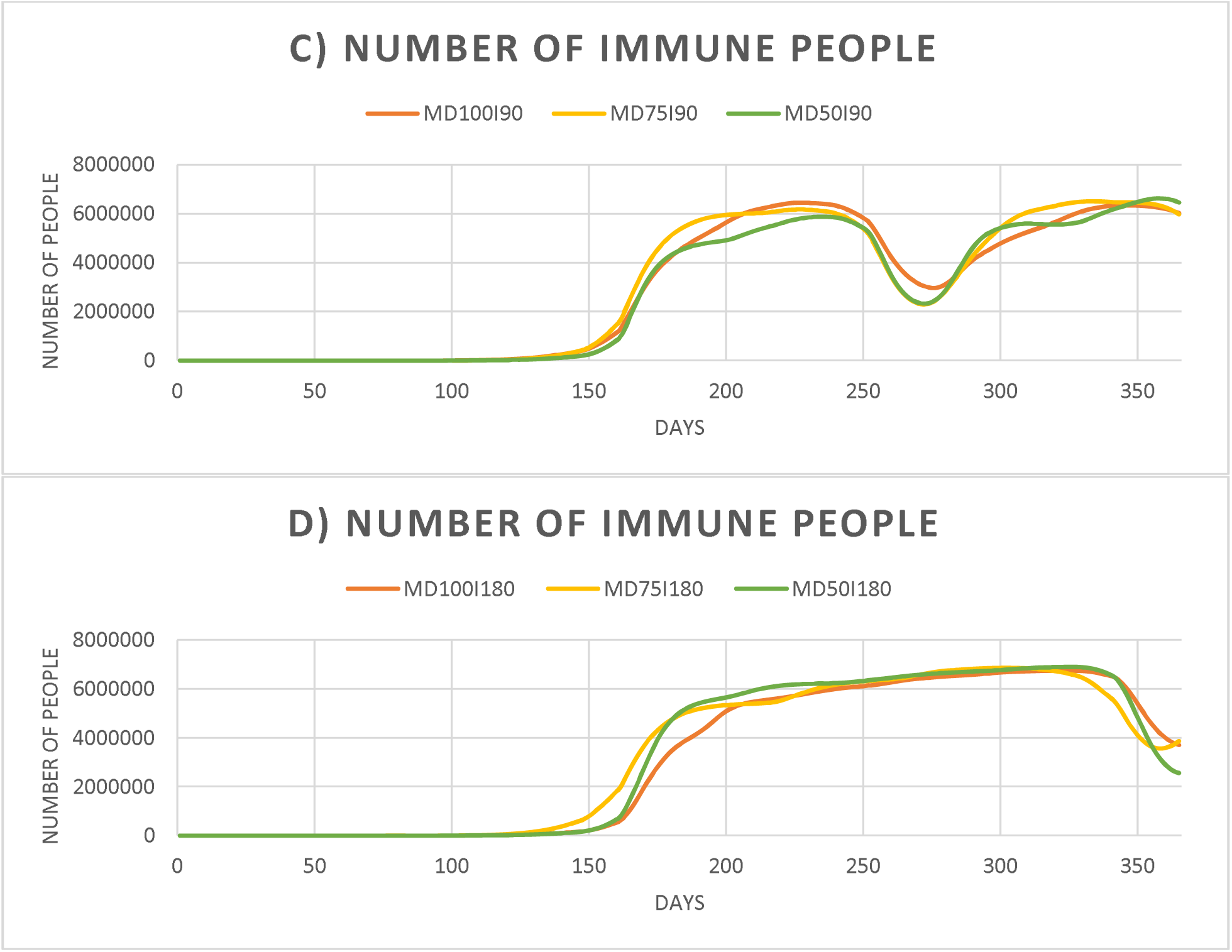
a): Number of Deceased people (90 days immunity scenarios); b) Number of Deceased people (180 days immunity scenarios); c): Number of Immune people (90 days immunity scenarios); b) Number of Immune people (180 days immunity scenarios)

The trends in which the curves of figures 5 a), b), and 6 a), b), c) and d) move are all similar with a time-offset denoting the time in which people stay in each of the intermediate states. Though most of the patients recover, major proportion of people being asymptomatic remains a great challenge for contact tracing and isolation. To measure the protection offered by the lockdown and use of control measures, Relative Risk (RR), Attributable Risk (AR), Population Attributable Risk (PAR) and PAR % were determined for the various scenarios (table 4). The first time period of 104 days indicates the time period after which the first recovered person would lose immunity. Successively, these parameters are calculated for further time periods to analyze how they vary for different lockdown scenarios [35; 41].

**Figure 6.**
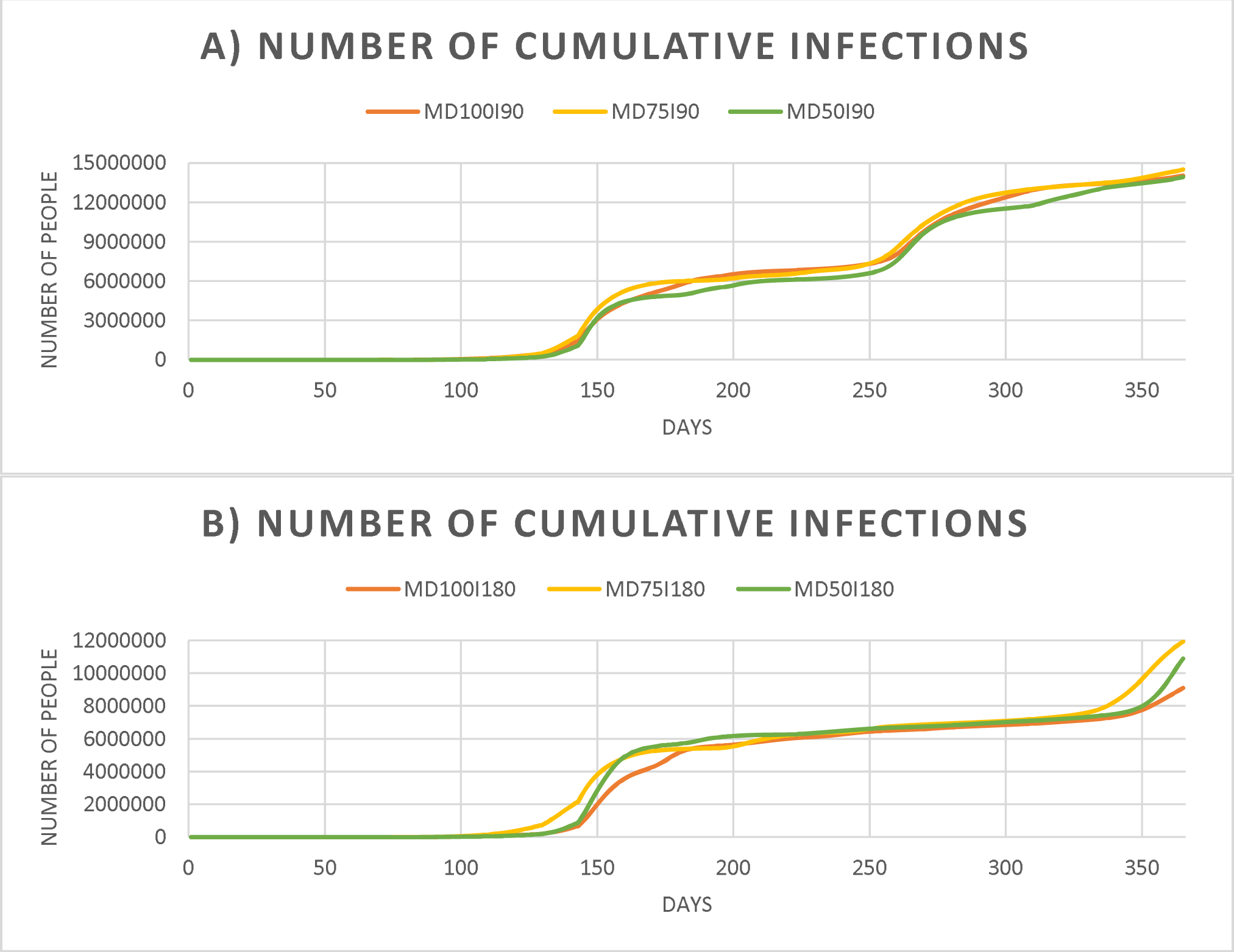
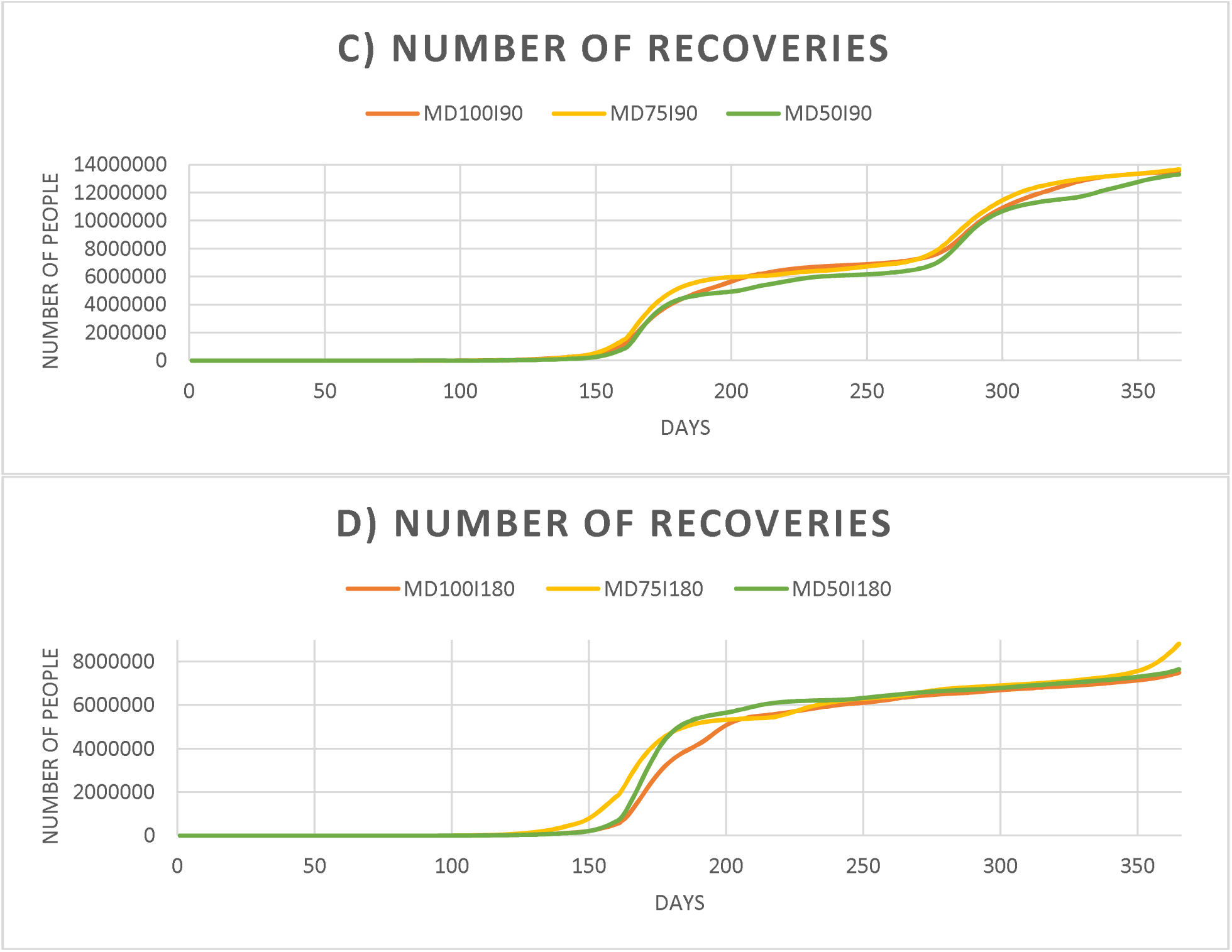
a): Number of Cumulative infections (90 days immunity scenarios); b) Number of Cumulative infections (180 days immunity scenarios); c): Number of Recoveries (90 days immunity scenarios); b) Number of Recoveries (180 days immunity scenarios)

**Figure 7:**
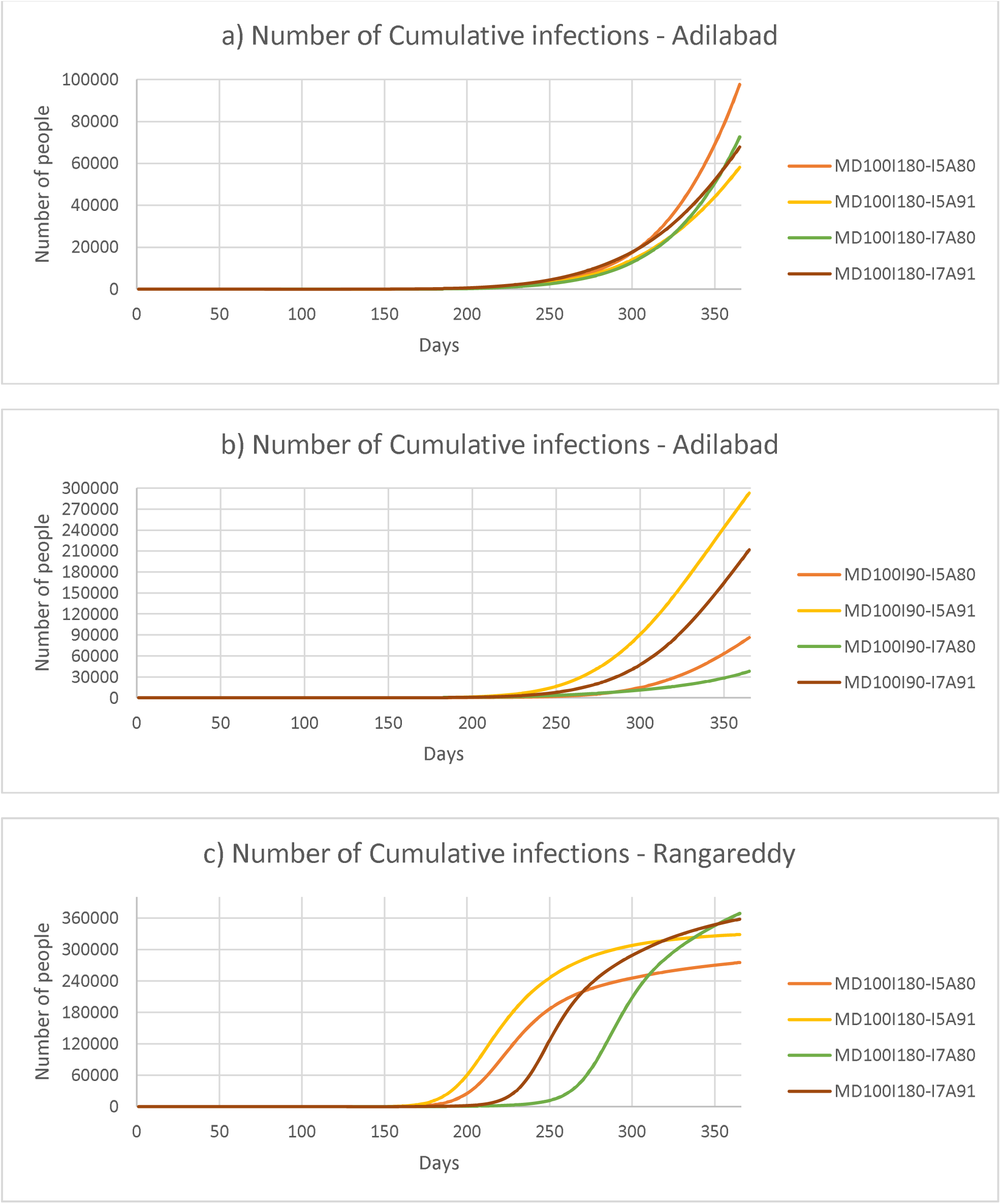
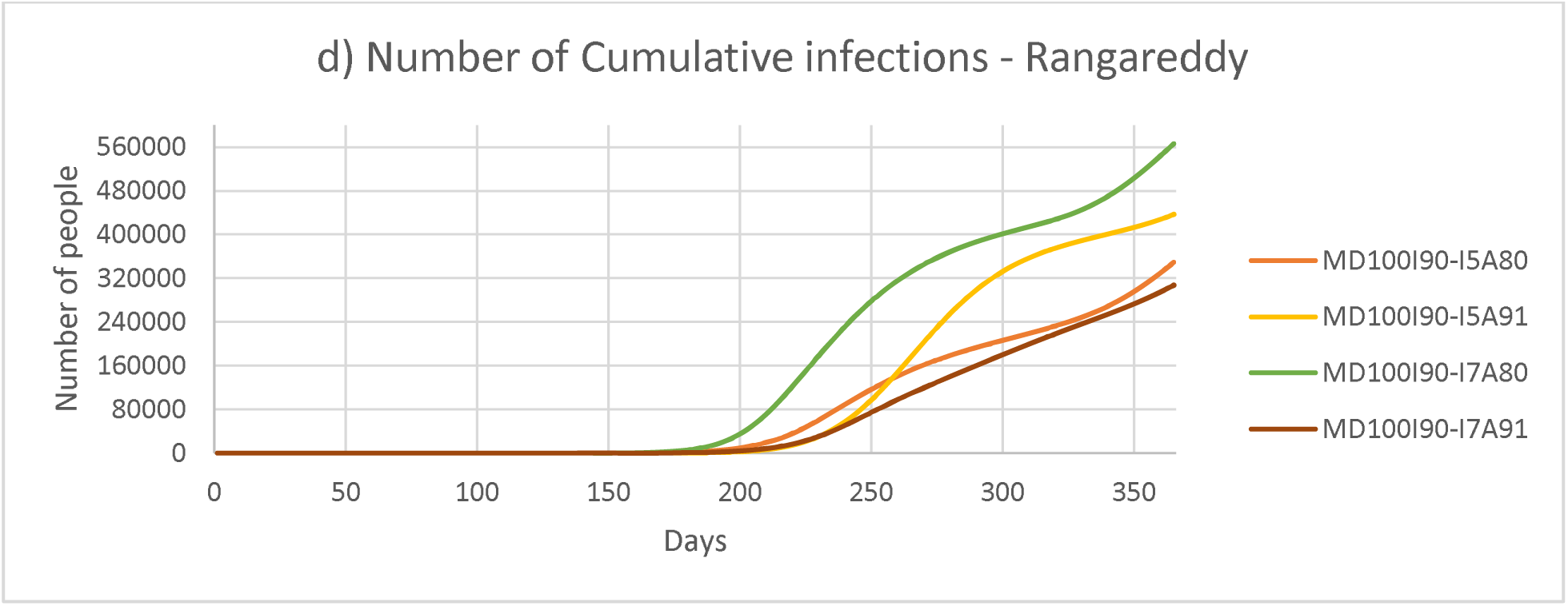
a) Number of Cumulative infections - Adilabad, b) Number of Cumulative infections - Adilabad, c) Number of Cumulative infections - Rangareddy, d) Number of Cumulative infections - Rangareddy

## 4. Sensitivity Analysis

The dynamics of pandemic that has been changing consistently is making the scenario challenging for the healthcare fraternity and policymakers. In India, recent studies have reported prolonged incubation period with a mean of 6.93 days [59] and the proportion of asymptomatic infections to be 91 percent [45]. Sensitivity analysis has been conducted to examine the effects of these changes on the disease transmission. Various scenarios of sensitivity analysis contain notations in which the numbers succeeding ‘I’ and ‘A’ indicate incubation period (days) and the proportion of asymptomatic people respectively.

Results in table 6 depict that the spread is relatively higher in Rangareddy compared to Adilabad, which is due to the higher population density of the district. Cumulative infections in most of the scenarios are higher when the incubation period is 7 days. This is because of the additional days in which infected people transmit infection. Also, in scenarios wherein asymptomatic proportion is considered to be 91 percent, the admissions are less because symptomatic people are only considered to be admitted whilst asymptomatic people are assumed to recover without being treated in hospitals. More asymptomatic carriers highly increase the spread of infection. Increasing testing, contact tracing, etc., and pitching in NPIs to track the close contacts of infected people might help to curtail the infection. The results of figure 6 also complements the fact that as the asymptomatic proportion and/ or incubation period increase, the spread of infection increases

**Table 5:**
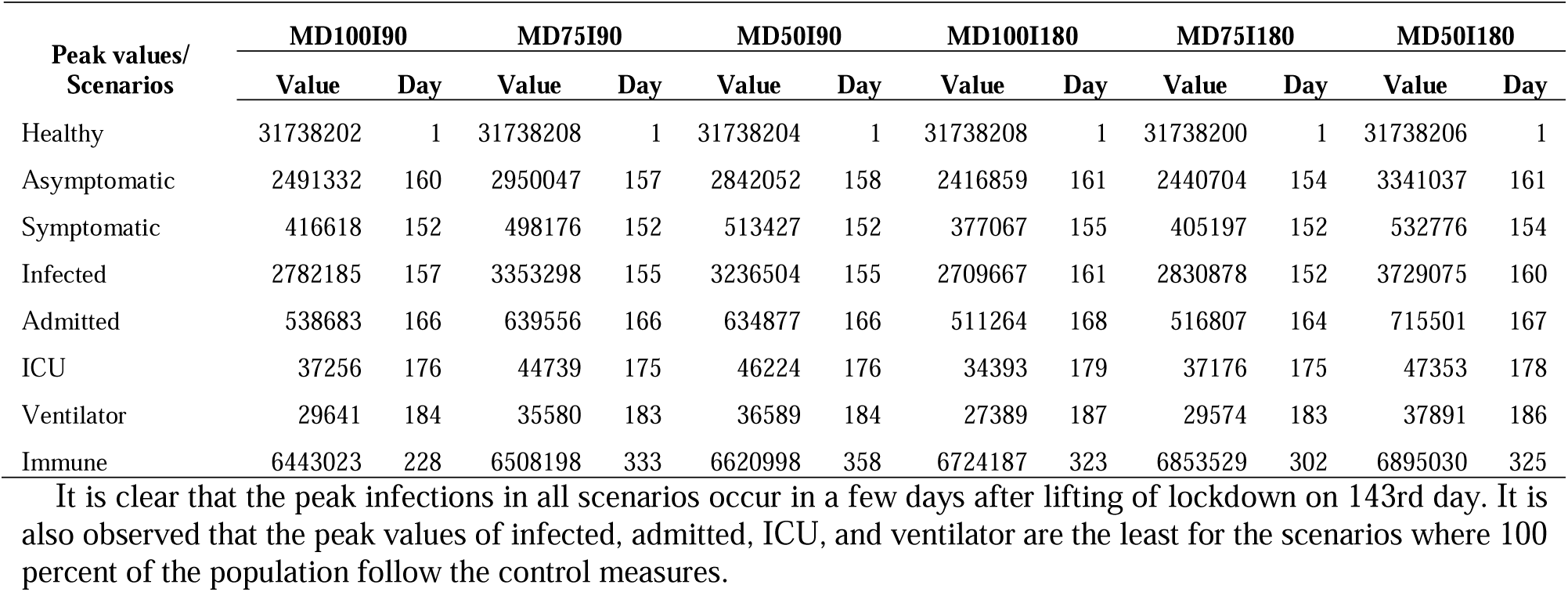
Peak values

**Table 6:**
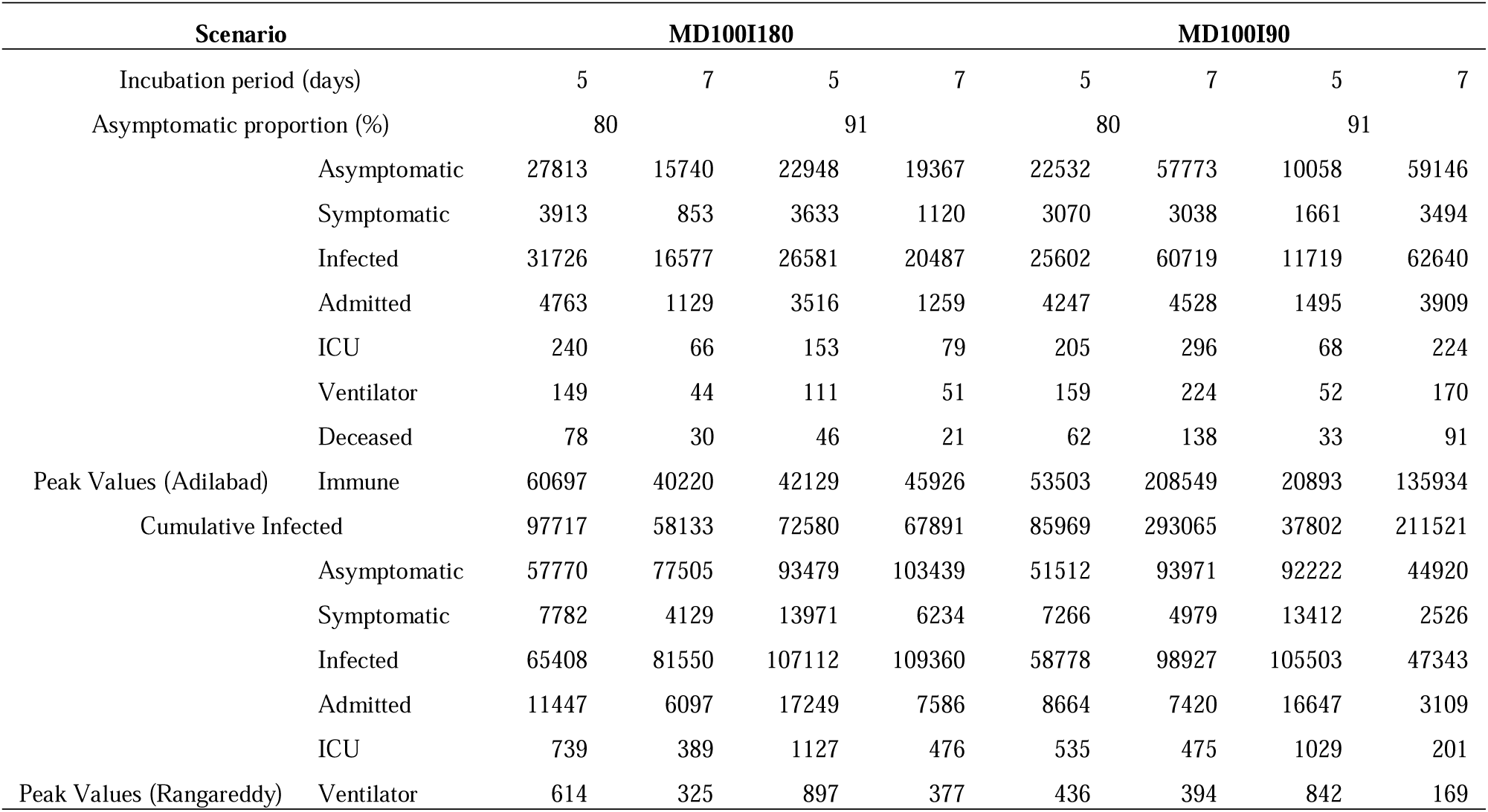
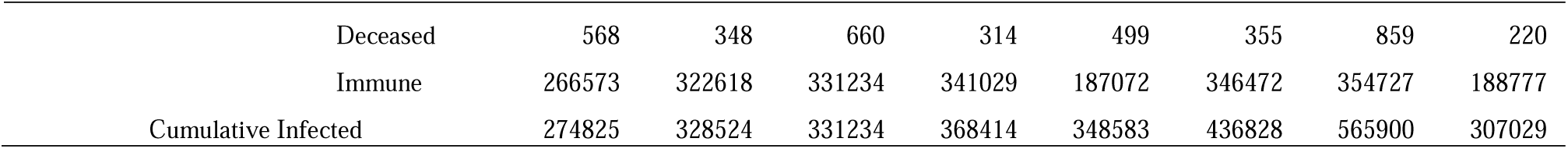
Results of Sensitivity Analysis

## 5. Discussion

The present study simulated 31738240 valid agents representing 90.67% of Telangana population for six different scenarios considering the various phases of lockdown as was imposed in India. The study also measures the effect of use of control measures and role of immunity in the spread of infection. This places policymakers in a better position to take decisions locally [10]. District level parameters have been considered to run the simulation along with assignment of agent-level details such as age, geospatial locations, etc. The model description, results, and discussion are inline with ethical good practice in modelling and ISPOR-SMDM Modeling Good Research Practices.

Present study involves modeling of six different scenarios with varying proportions of people using control measures (100%, 75% and 50%) and varying immunity periods of recovered patients (90 and 180 days). Likewise, several other studies have modeled different types of scenarios. Most of the studies revolve around imposing lockdowns whilst varying the durations of lockdown [37], isolating vulnerable population [37], varying lockdown stringency based on age [36], changing proportions of contacts made outside household and close contacts [40], considering contact tracing measures [36; 68], etc.

In this study, we have developed the agents from the 2011 Census of India data with 31738240 valid agents representing 90.67 percent of Telangana’s population [57; 70]. Number of people in each of the states have been determined based on parameters of each district as localized models are much preferred for decision-making. Synthetic population approach that is widely employed use open data sources like the Australian Census data [64], US Census data [36; 40], London Imperial College data [37], etc. Various geolocations have been represented previously using different number of agents such as 5000 agents in a University of Italy [21], 500000 agents of NYC [36], 24 million agents of entire Australian population [64], 750805 agents of Urmia, Iran [51], 10 million agents of Delaware, US [40], etc.

Contact network influences the behavior of agents in the network. Present study has considered contacts made in closest circle and external places as mentioned in table 3. As the stringency of lockdown is increased, the overall contact rate decreases whereas the proportion of contacts made in closer circle and households increase. Different studies have considered a range of contact network settings such as contacts in closed environments as offices, colleges, contacts based on schedules [36; 40; 68], indirect transmission through viral particles [40], touching of contaminated objects, enhancing protection by washing hands [20; 43; 68], inclusion of travel medium and routes [27; 60], transmission in public places [40], etc.

From the results, it is clear that the rate of transmission of infection increased as the lockdowns were lifted and the peak infections were observed in a lesser number of days post-lockdown. Also, the values of RR, AR, PAR% indicate that the protection factor had higher values during lockdown and when a higher number of people followed control measures. The values of AR, RR and PAR% help in determining the protection offered, strength, and percent of population that could attribute to the protection factor [41]. Effect of immunity also provides information about possible secondary infections post loss of immunity. This assists the capacity planning of health care practitioners and policymakers. Complementing this is the study by The Center For Disease Dynamics, Economics & Policy (CDDEP) and Princeton University where they have provided state-wise estimates across India to assist policymakers to cope up with the influx of infections [42; 73].

Limitations to the study include the exclusion of comorbidities among patients, transportation modes, indirect transmission through suspended particles, etc., which could be considered to improve the accuracy of the model. Considering more parameters are however limited to the availability and authenticity of data.

## 6. Conclusions

We have modeled the COVID-19 transmission dynamics considering the various lockdown phases of India using an ABM approach using Python, an open source coding platform. Localized studies such as this, based on synthetic populations could be helpful in decision-making processes of localized authorities. Important factors such as protective factor could provide insights on the proportion of population that would be shielded by imposing control measures.

## Data Availability

Python code and detailed resutls have all been provided in the below link.

https://osf.io/3nxby/?view_only=96320e1dd7f048318294898ccd657275

## Data Availability

The python code, and detailed district-wise estimates files are available in the link: https://osf.io/3nxby/?view_only=96320e1dd7f048318294898ccd657275

